# A Normative Database of A-Scan Data Using the Heidelberg Spectralis Spectral Domain Optical Coherence Tomography Machine

**DOI:** 10.1101/2021.02.16.21251860

**Authors:** Joos Meyer, Roshan Karri, Helen Danesh-Meyer, Kate Drummond, Andrew Symons

## Abstract

**Purpose:** Develop the first normative database of macular and circumpapillary scans with reference values at the level of the A-scan using the Heidelberg Spectralis Optical Coherence Tomography (OCT) machine.

**Methods:** This study is a retrospective cross sectional analysis of macular and circumpapillary OCT scans of healthy individuals. All participants had a full ophthalmic examination, including best corrected visual acuity, intraocular pressure, biomicroscopy, posterior segment examination and OCT scan. The volume and thickness of each of the nine Early Treatment Diabetic Retinopathy zones at the macula were analysed for the total retinal thickness, retinal nerve fibre layer (RNFL), ganglion cell layer (GCL) and inner plexiform layer (IPL). The thickness of the circumpapillary RNFL was analysed at the disc. De-identified A-scans were extracted from the OCT machine as separate tab-separated text file and made available according to the data sharing statement.

**Results:** Two-hundred eyes from 144 participants were included of which 98 (49%) were female. The mean age (SD) was 48.52 (17.52). Participants were evenly distributed across four age groups and represented nine broad ethnic groups in proportions comparable to the local distribution. All the macular scans were 20° x 20° (5.9 mm x 5.9 mm), with a total scan density between 12,800 and 49,152 A-scans. The peripapillary scans were all 12° (3.5 mm), at a scan density of 768 A-scans. The mean retinal, GCL and IPL volumes were significantly greater in males than females. Age and total retinal volume (r = –0.2561), GCL volume (– 0.2911) and IPL volume (–0.3194) were negatively correlated. No significant correlation was found between the RNFL and age.

**Conclusion:** This study provides a normative database of macular and circumpapillary scans with reference values at the level of the A-scan using the Heidelberg Spectralis Optical Coherence Tomography (OCT) machine.

## Introduction

Optical coherence tomography (OCT) is a non-invasive imaging technology that uses low coherence interferometry to create two-dimensional cross-sectional imaging of biological systems.[1] In ophthalmic practice, OCT is used to image and measure the retinal architecture and changes that occur in disease states. Diseases, such as glaucoma and compressive optic neuropathy, manifest distinct changes on OCT scans, particularly on the inner retinal layers. Important OCT measurements for both diseases include the circumpapillary retinal nerve fibre layer (RNFL), macular RNFL and the macular ganglion cell layer (GCL).[2,3]

The Early Treatment Diabetic Retinopathy Study (ETDRS) grid is a standardised pattern of dividing and measuring the thickness and volume profile of the retina.[4] It was first introduced in 1980, and is still the primary method of reporting.[5] Since its inception, additional grid patterns have been developed; however, there is limited flexibility for clinicians and researchers to explore novel patterns of segmentation within any currently commercially available software.

For the macular, the standard 20° x 20° scan consists of horizontally stacked scan lines to create a square scan area. Each horizontal line is termed a B-scan, and each of these comprises a row of equally distanced A-scans. The A-scan represents the most basic unit of interrogation from which the rest of the scan is generated. The scan density of the Heidelberg Stratus (Heidelberg Engineering, Heidelberg, Germany) varies between 25,000 and 50,000 A-scans. For the optic disc, the standard circumpapillary scan consists of a 360° radial scan of 720 A-scans. Extracting the A-scan data enables the exploration of OCT changes that do not conform to the spatial distribution of the predefined grid patterns.

A normative database of scans is needed for comparison to validate changes in disease states. To the author’s knowledge, a normative database that contains both macular and circumpapillary scans, which includes reference values for each A-scan of the Heidelberg Stratus, does not currently exist.

The purpose of this study is to report the normative values for macular and optic disc scans within the standard ETDRS grid and at the level of the A-scan. This study aims to help clinicians and researchers to identify patterns of change that occur outside the standard segmentation patterns of the retina.

## Methods

### Study Population

This study is a retrospective cross-sectional analysis of 200 normal eyes that had a visual assessment and an OCT scan. The participants included patients of The Royal Melbourne Hospital (RMH) who were found to have no ocular pathology or systemic disease that could manifest in retinal changes. The RMH database was used to create this normative database. Approximately 50,000 eyes have been scanned since 2012 on the Heidelberg Spectralis machine at The RMH. The database was queried for normal OCT scans, and the potential participant’s medical records were checked against inclusion and exclusion criteria. All of the participants had a full ophthalmic examination, including their Snellen best corrected visual acuity (BCVA), intraocular pressure (IOP), biomicroscopy, posterior segment examination and OCT scan. The participant’s medical records were examined for any exclusions.

The retrospective inclusion criteria included healthy participants, aged 18 to 88, with a BCVA of 6/12 (20/40) on the Snellen chart measured at 6 m (20 feet), and high quality macular and peripapillary OCT scans (signal strength >15dB). The exclusion criteria included a history of any: (i) anterior segment disease (except cataracts); (ii) retinal disease; (iii) glaucoma; (iv) uveitis; (v) optic nerve disease; (vi) systemic disease known to affect the retina (e.g., diabetes); (vii) neurological disease; (viii) pituitary disease; (ix) intracranial space-occupying lesions; (x) previous neurosurgery; (xi) retinal lasers; (xii) vitrectomy; (xiii) topical or systemic anti-vascular endothelial growth factor treatment; (xix) IOP lowering medications; (xx) chemotherapy; or (xxi) previous systemic or active topical steroid use. The excluding biomarkers included peripapillary atrophy, RNFL haemorrhage, optic disc notching or thinning, age-related macular degeneration (including drusen) or macular disease on OCT scans. Ethnicity data were collected from the participants and categorised according to the Australian Bureau of Statistics’ (ABS) Australian Standard Classification of Cultural and Ethnic Groups (ASCCEG).[6]

This study was approved by the Human Research and Ethics Committee of The RMH. It was conducted according to the Declaration of Helsinki in its currently applicable version.

### Optical Coherence Tomography

All the OCT scans were performed with the Heidelberg Spectralis Spectral Domain (SD) OCT and the Heidelberg Eye Explorer version 6.7.13.0 (Heidelberg Engineering, Heidelberg, Germany). All scans were completed by experienced medical imaging personnel at The RMH medical imaging department; they were performed in a dark room as per the standard hospital protocol. Only the well-centred scans (>15dB quality) were included in the analysis. The number of dioptres focus spherical was recorded for each scan as a proxy for the refractive error. The scans were quality controlled for the accurate segmentation of each macular retinal inner layer and peripapillary scan, and an experienced grader manually corrected any aberrations.

The macular scans were divided into 1 mm, 3 mm and 6 mm rings on the macular ETDRS map. The inner ring was defined as the central thickness, and the middle and outer rings were divided into four zones designated as the superior, nasal, inferior and temporal zones. The average thickness in each of the nine zones, the macular thickness and the full 360° peripapillary scans were included in the final analysis.

### A-Scan Data

The A-scans were extracted from the machine using the Heidelberg Spectralis Layer Segmentation Export Special Function version 6.0. Each file contained the results from the automatic retinal layer segmentation algorithm of a Spectralis OCT machine as a tab-separated text file. The distance values of each A-scan location were represented as relative coordinates in nanometres from the probe to the layer segmentation interface. The normative values of the A-scans were compiled as a de-identified single data array per patient, per eye.

### Statistical Analysis

All analyses were conducted in the R statistical programming language using RStudio version 1.3.1073 (Boston, USA).[7] The exploratory data analysis and data visualisation were performed using the ggplot2 package.[8] The descriptive statistics were reported as the mean, standard deviation and the first, fifth and 95th percentiles. An independent t-test was used to compare the thickness values between groups. The correlation between measurements was found using Pearson correlation coefficients. A multivariate analysis was used to consider the effects of age and gender; an alpha of 0.05 was considered significant. The chi-square goodness of fit test was used to compare the observed distribution of ABS ethnicities to the expected distribution in Melbourne, Australia.

## Results

### Study participants

Two-hundred eyes from 146 participants were included in the final analysis. There were 69 (47%) females and 77 (53%) males, and the mean age (SD) was 48.52 (17.52) (range: 21–85). Mean (SD) scan focus was 0.29 D (1.96) (range: −6.48–7.86). The participants were evenly distributed across the age groups (Table 1). Females were slightly under-represented in the younger age groups and over-represented in the older age groups. This study included 101 (50.5%) right eyes and 99 (49.5%) left eyes. Participants from nine broad ethnicity groups were included in the study (Table 2). There were significant differences in the proportion of broad ethnic categories compared to the census population proportions in four out of the nine groups.

**Table 1.**
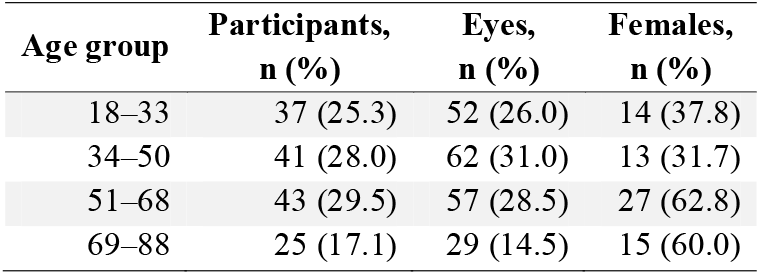
Distribution of participants by age group and gender.

**Table 2.** Australian Bureau of Statistics Australian Standard Classification of Cultural and Ethnic Groups broad group classification of participants.

| Broad ethnicity category | Participants, Melbourne | P |
| --- | --- | --- |

|  | n (%) | (%) |  |
| --- | --- | --- | --- |
| North African and Middle Eastern | 15 (7.5) | 1.9 | <0.001 |
| Northeast Asian | 4 (2) | 5.1 | 0.0644 |
| Northwest European | 9 (4.5) | 5.3 | 0.7283 |
| Oceanian | 111 (55.5) | 68.5 | <0.001 |
| People of the Americas | 2 (1) | 0.8 | 1 |
| Southeast Asian | 14 (7) | 6.0 | 0.644 |
| Southern and Central Asian | 22 (11) | 6.9 | 0.031 |
| Southern and Eastern European | 15 (7.5) | 4.5 | 0.0666 |
| Sub Saharan African | 8 (4) | 1.0 | <0.001 |

### A-scans

Two-hundred de-identified tab separated text files were exported from the Heidelberg Stratus SD OCT machine. All the macular scans were 20° x 20° (5.9 mm x 5.9 mm), with 25 to 96 B-scans, and 512 A-scans per B-scan with the automatic real-time mode active. The total scan density was between 12,800 and 49,152 A-scans. The peripapillary scans were all 12° (3.5 mm diameter), at a scan density of 768 A-scans. All the scan data are available, according to the data sharing statement.

### Distribution of macular and circumpapillary measurements

The mean (SD) total retinal volume was 8.67 mm^3^ (0.38) (range: 7.65–9.58). The macular RNFL volume (SD) was 0.96 mm^3^ (0.11) (range: 0.72–1.26). The GCL volume (SD) was mm^3^ 1.09 (0.10) (range: 0.82–1.31) and the inner plexiform layer (IPL) volume (SD) was 0.89 mm^3^ (0.07) (range: 0.7–1.05). See Figs 1 and 2 demonstrating layer measurement distributions.

**Fig 1.**
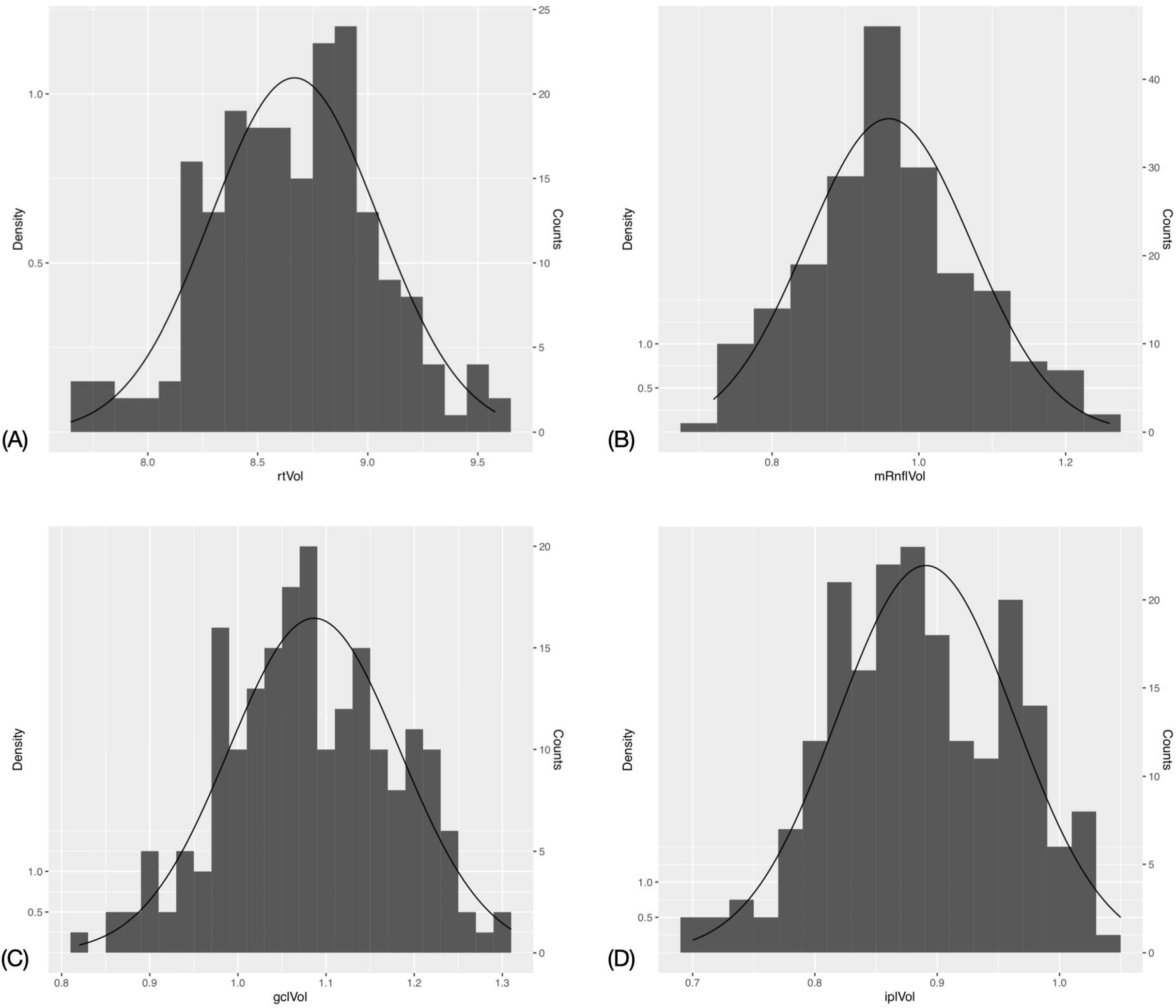
Histogram and fitted normal distribution curves of macular optical coherence tomography measurements. (A) Total retinal volume. (B) Retinal nerve fibre layer volume. (C) Ganglion cell layer volume. (D) Inner plexiform layer volume.

**Fig 2.**
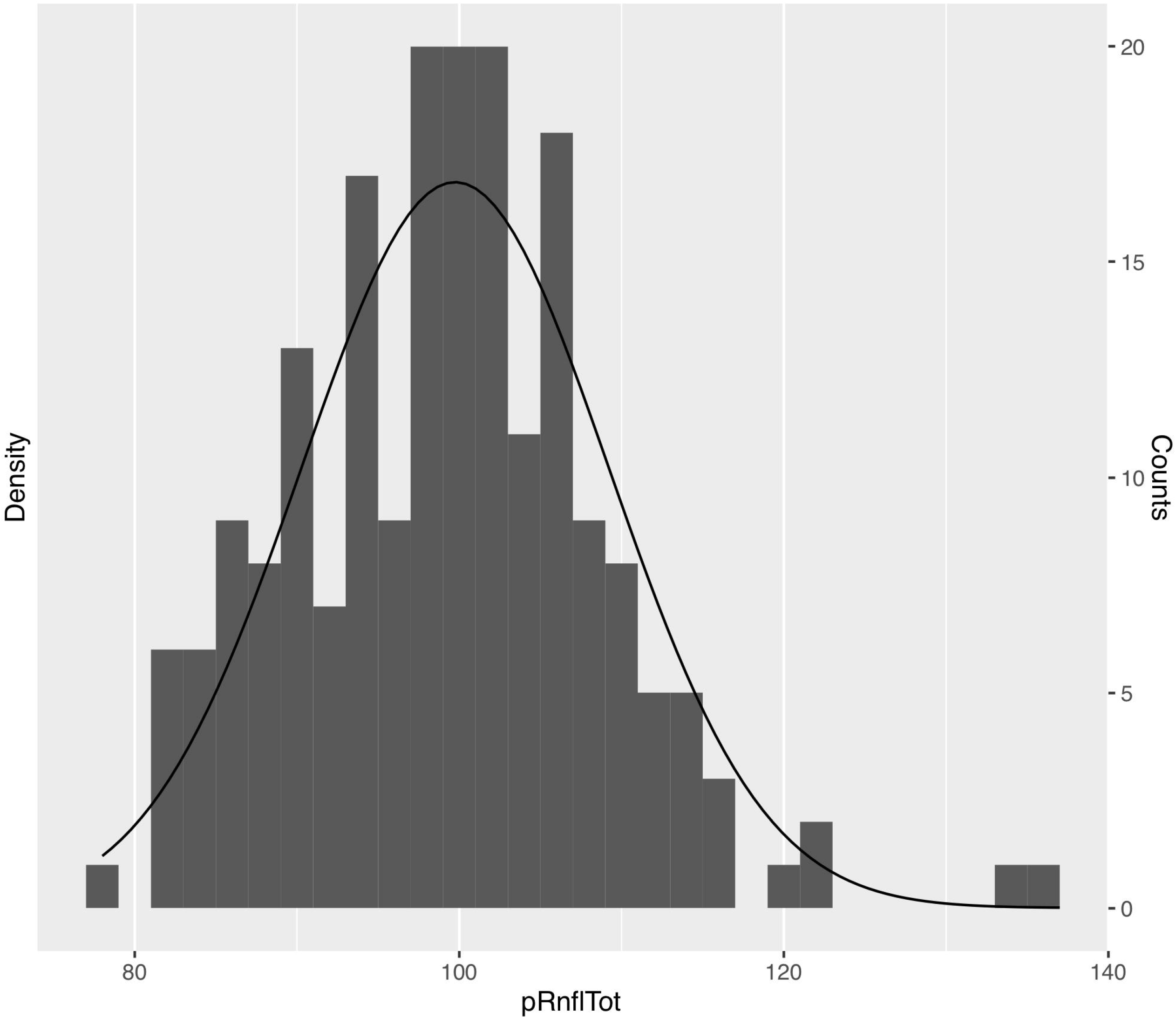
Histogram and fitted normal distribution curve of mean peripapillary retinal nerve fibre layer thickness values.

### Difference in measurements between males and females

The multivariate regression analysis controlling for age and ethnicity (Table 3) showed that the retinal and GCL volumes were significantly less for females. The total RT was significantly lower in females in all sectors except for the superior outer sector. The same was true for the central, nasal inner, temporal inner, inferior inner macular RNFL, GCL and IPL thicknesses. The GCL and IPL also showed that females had significantly lower superior inner sectors. There were no significant differences found at the peripapillary RNFL between males and females (Table 4).

**Table 3.** Female regression coefficient in multivariate analysis controlling for age with macular sectorial thickness values as outcome.

| Sector | Retina | p | mRNFL | p | GCL | p | IPL | p |
| --- | --- | --- | --- | --- | --- | --- | --- | --- |
| Volume | -0.175751 | *** | -0.0112461 |  | -0.0309332 | * | -0.0121464 |  |
| Centre | -14.58119 | *** | -1.287385 | *** | -2.741456 | *** | -2.84705 | *** |
| Nasal inner | -11.15093 | *** | -0.86165 | * | -2.44177 | ** | -1.23396 | * |
| Nasal outer | -5.82186 | ** | -0.602074 |  | -0.34252 |  | 0.09550 |  |
| Superior inner | -10.07472 | *** | -0.92011 |  | -1.82976 | ** | -1.00703 | * |
| Superior outer | -2.88614 |  | 0.97539 |  | -0.51977 |  | 0.02971 |  |
| Temporal inner | -10.548601 | *** | -0.503470 | * | -3.26108 | *** | -1.68946 | ** |
| Temporal outer | -4.83874 | * | -0.39463 |  | -1.01023 |  | -0.97755 |  |
| Inferior inner | -10.35359 | *** | -1.438830 | ** | -1.97374 | ** | -1.07487 | * |
| Inferior outer | -4.99137 | * | -0.33538 |  | -0.37526 |  | -0.13309 |  |

P-values are coded as: \*\*\*<0.001, \*\*<0.01 and \*<0.05.

**Table 4.** Female regression coefficient in multivariate analysis controlling for age with peripapillary retinal nerve fibre layer sectoral thickness values as outcome.

| Sector | Disc | p |
| --- | --- | --- |
| Total | -0.58076 |  |
| Nasal superior | 2.71149 |  |
| Nasal | -0.18586 |  |
| Nasal inferior | -0.08341 |  |
| Temporal superior | -2.02026 |  |
| Temporal | -0.84995 |  |
| Temporal inferior | -2.00331 |  |

P-values are coded as: \*\*\*<0.001, \*\*<0.01 and \*<0.05.

### Decrease in thickness per decade

The mean retinal, GCL and IPL volumes were found to be significantly greater in males than females (Table 5). The mean volumes, ETDRS segment thickness values and the first, fifth and 95th percentile values were grouped by age for each macular layer (Table 6-9). The mean (SD) peripapillary RNFL thickness was 99.7 μm (9.47) (range: 78–137). The mean peripapillary RNFL thicknesses were grouped by age (Table 10).

**Table 5.** Mean volumes by gender.

| Volume | Male | Female | P |
| --- | --- | --- | --- |
| Retinal | 8.77 (0.401) | 8.56 (0.326) | >0.0001 |
| RNFL | 0.961 (0.122) | 0.957 (0.101) | 0.7943 |
| GCL | 1.11 (0.11) | 1.07 (0.0764) | 0.002346 |
| IPL | 0.903 (0.0801) | 0.879 (0.0622) | 0.01882 |
| Disc RNFL thickness | 99.9 (9.66) | 99.6 (9.31) | 0.8 |

**Table 6.** Macular retinal thickness distribution by age group, showing mean, first, fifth and 95th percentile values for each segment.

| Retina | 18–33 |  |  |  | 34–50 |  |  |  | 51–68 |  |  |  | 69–88 |  |  |  |
| --- | --- | --- | --- | --- | --- | --- | --- | --- | --- | --- | --- | --- | --- | --- | --- | --- |
|  | Percentile |  |  |  | Percentile |  |  |  | Percentile |  |  |  | Percentile |  |  |  |
|  | x□ | 1st | 5th | 95th | x□ | 1st | 5th | 95th | x□ | 1st | 5th | 95th | x□ | 1st | 5th | 95th |
| Volume | 8.74 | 7.99 | 8.11 | 9.13 | 8.76 | 7.77 | 8.22 | 9.27 | 8.59 | 8.12 | 8.16 | 9.45 | 8.48 | 7.66 | 7.71 | 9.02 |
| Centre | 270.56 | 228.5 | 239.3 | 306.8 | 280.50 | 228.6 | 246.1 | 308.0 | 279.05 | 220.7 | 237.0 | 320.2 | 270.17 | 233.0 | 240.0 | 305.4 |
| Nasal inner | 348.29 | 321.0 | 327.0 | 369.7 | 353.31 | 314.0 | 329.0 | 378.0 | 345.77 | 319.7 | 324.2 | 379.2 | 342.45 | 315.6 | 317.4 | 371.6 |
| Nasal outer | 321.37 | 294.6 | 298.1 | 341.9 | 319.76 | 280.4 | 298.1 | 338.0 | 312.86 | 283.0 | 293.6 | 343.6 | 308.34 | 278.6 | 280.0 | 333.6 |
| Superior inner | 343.29 | 254.7 | 320.1 | 368.3 | 351.45 | 310.1 | 334.1 | 375.0 | 342.33 | 312.0 | 320.4 | 369.2 | 339.69 | 308.6 | 311.2 | 365.8 |
| Superior outer | 302.37 | 272.0 | 276.7 | 321.8 | 301.40 | 269.5 | 285.0 | 320.0 | 296.14 | 273.7 | 278.8 | 322.6 | 291.38 | 259.1 | 262.8 | 310.6 |
| Temporal inner | 329.56 | 300.0 | 305.1 | 352.9 | 337.15 | 295.6 | 318.0 | 360.9 | 330.67 | 302.2 | 304.8 | 362.6 | 327.41 | 299.1 | 302.0 | 346.2 |
| Temporal outer | 286.73 | 257.5 | 267.1 | 300.5 | 286.18 | 251.7 | 263.1 | 304.0 | 282.26 | 263.0 | 265.8 | 315.0 | 278.62 | 244.1 | 247.8 | 299.2 |
| Inferior inner | 343.35 | 315.1 | 320.2 | 362.9 | 347.48 | 309.6 | 328.0 | 370.9 | 339.61 | 316.0 | 316.8 | 371.0 | 338.31 | 309.4 | 314.2 | 361.2 |
| Inferior outer | 292.69 | 258.1 | 266.1 | 309.4 | 291.69 | 266.8 | 272.0 | 308.0 | 286.37 | 264.1 | 268.4 | 314.2 | 282.48 | 253.3 | 255.6 | 303.2 |

**Table 7.** Macular retinal nerve fibre layer thickness distribution by age group, showing mean, first, fifth and 95th percentile values for each segment.

| RNFL | 18–33 |  |  |  | 34–50 |  |  |  | 51–68 |  |  |  | 69–88 |  |  |  |
| --- | --- | --- | --- | --- | --- | --- | --- | --- | --- | --- | --- | --- | --- | --- | --- | --- |
|  | Percentile |  |  |  | Percentile |  |  |  | Percentile |  |  |  | Percentile |  |  |  |
|  | x□ | 1st | 5th | 95th | x□ | 1st | 5th | 95th | x□ | 1st | 5th | 95th | x□ | 1st | 5th | 95th |
| Volume | 0.95 | 0.77 | 0.81 | 1.12 | 0.96 | 0.74 | 0.76 | 1.16 | 0.94 | 0.75 | 0.78 | 1.11 | 1.01 | 0.79 | 0.88 | 1.16 |
| Centre | 12.42 | 8.0 | 9.0 | 16.0 | 12.98 | 6.6 | 10.0 | 16.0 | 12.91 | 8.6 | 9.0 | 16.2 | 12.41 | 8.0 | 8.8 | 15.0 |
| Nasal inner | 22.06 | 17.5 | 19.0 | 26.0 | 22.31 | 17.0 | 18.0 | 27.0 | 21.96 | 17.0 | 18.0 | 27.0 | 22.69 | 18.0 | 18.4 | 28.0 |
| Nasal outer | 51.73 | 39.0 | 43.0 | 65.0 | 51.44 | 38.0 | 38.0 | 69.9 | 48.79 | 29.1 | 38.8 | 58.8 | 53.24 | 25.5 | 40.6 | 64.0 |
| Superior inner | 25.54 | 19.5 | 21.6 | 31.4 | 26.02 | 19.6 | 20.1 | 32.0 | 25.28 | 19.6 | 20.8 | 31.2 | 27.45 | 20.8 | 23.0 | 35.2 |
| Superior outer | 38.15 | 27.5 | 31.1 | 45.0 | 39.26 | 28.6 | 30.1 | 50.8 | 37.68 | 26.0 | 28.6 | 45.6 | 41.62 | 30.4 | 34.4 | 50.8 |
| Temporal inner | 17.56 | 15.0 | 16.0 | 19.0 | 17.90 | 16.0 | 16.0 | 20.0 | 18.37 | 16.0 | 16.0 | 21.2 | 18.38 | 16.3 | 17.0 | 20.6 |
| Temporal outer | 18.73 | 16.0 | 17.0 | 20.0 | 18.77 | 16.6 | 17.0 | 21.0 | 20.39 | 16.6 | 17.0 | 23.0 | 21.07 | 19.0 | 19.0 | 23.0 |
| Inferior inner | 26.37 | 22.0 | 23.0 | 31.0 | 26.53 | 19.0 | 21.1 | 34.0 | 26.42 | 20.1 | 22.0 | 31.6 | 27.00 | 22.3 | 23.0 | 32.6 |
| Inferior outer | 42.27 | 30.0 | 32.0 | 49.0 | 41.50 | 30.2 | 32.0 | 53.0 | 41.79 | 30.0 | 31.8 | 53.2 | 42.72 | 33.0 | 33.4 | 51.2 |

**Table 8.**
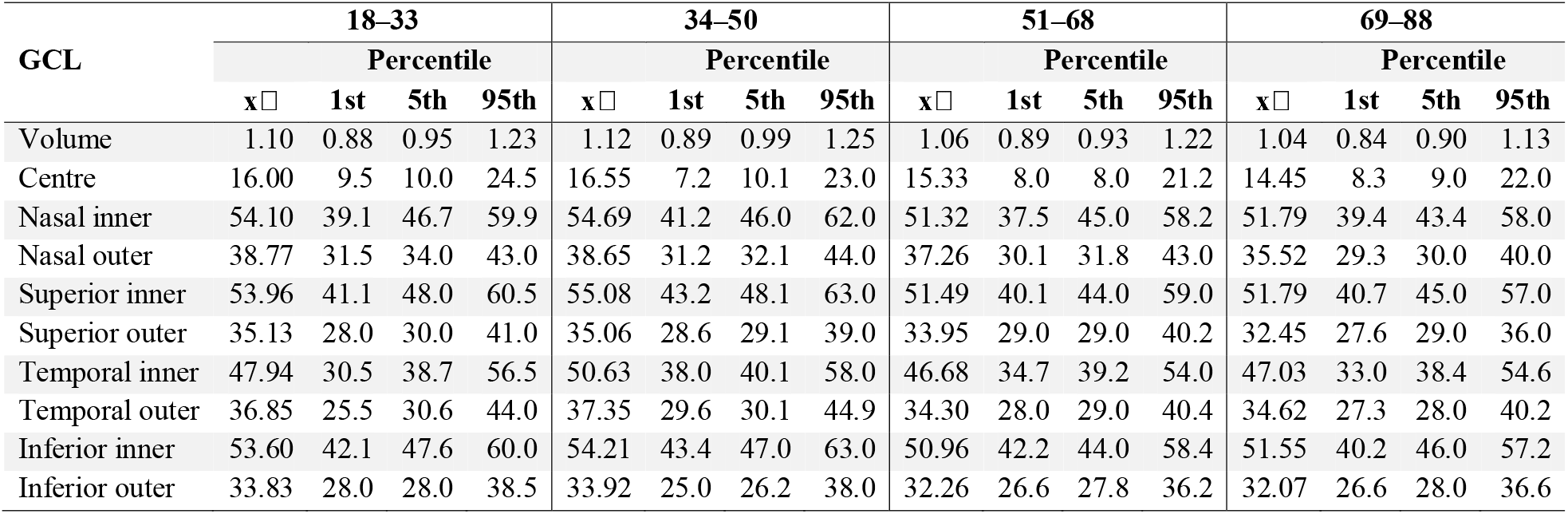
Macular ganglion cell layer thickness distribution by age group, showing mean, first, fifth and 95th percentile values for each segment.

**Table 9.** Macular inner plexiform layer thickness distribution by age group, showing mean, first, fifth and 95th percentile values for each segment.

| IPL | 18–33 |  |  |  | 34–50 |  |  |  | 51–68 |  |  |  | 69–88 |  |  |  |
| --- | --- | --- | --- | --- | --- | --- | --- | --- | --- | --- | --- | --- | --- | --- | --- | --- |
|  | Percentile |  |  |  | Percentile |  |  |  | Percentile |  |  |  | Percentile |  |  |  |
|  | x□ | 1st | 5th | 95th | x□ | 1st | 5th | 95th | x□ | 1st | 5th | 95th | x□ | 1st | 5th | 95th |
| Volume | 0.91 | 0.79 | 0.80 | 1.03 | 0.91 | 0.71 | 0.81 | 0.99 | 0.87 | 0.73 | 0.77 | 1.01 | 0.86 | 0.72 | 0.76 | 0.93 |
| Centre | 21.15 | 14.0 | 15.0 | 27.9 | 22.24 | 13.6 | 15.1 | 28.0 | 21.53 | 14.6 | 15.8 | 27.2 | 20.90 | 15.3 | 16.0 | 26.0 |
| Nasal inner | 44.12 | 34.5 | 39.0 | 50.0 | 44.35 | 35.6 | 38.1 | 50.0 | 42.21 | 35.1 | 37.0 | 48.0 | 42.07 | 33.4 | 37.4 | 46.0 |
| Nasal outer | 29.75 | 24.5 | 25.6 | 34.0 | 29.31 | 22.0 | 24.1 | 33.0 | 28.21 | 22.6 | 23.8 | 33.0 | 26.90 | 22.0 | 22.4 | 30.6 |
| Superior inner | 42.58 | 34.5 | 37.1 | 47.0 | 42.82 | 35.8 | 38.0 | 49.0 | 40.25 | 33.1 | 34.8 | 46.2 | 40.24 | 34.0 | 34.8 | 43.6 |
| Superior outer | 28.87 | 23.5 | 25.0 | 33.0 | 28.10 | 21.6 | 24.0 | 32.0 | 27.28 | 23.0 | 23.8 | 33.0 | 26.38 | 22.3 | 23.4 | 29.6 |
| Temporal inner | 42.17 | 30.6 | 35.6 | 48.5 | 43.16 | 34.6 | 38.0 | 49.0 | 41.16 | 33.7 | 35.8 | 47.2 | 41.07 | 32.1 | 35.8 | 45.0 |
| Temporal outer | 32.52 | 26.5 | 27.6 | 36.5 | 32.44 | 24.2 | 27.0 | 37.0 | 30.77 | 25.0 | 26.8 | 36.2 | 30.79 | 26.3 | 27.0 | 34.6 |
| Inferior inner | 42.46 | 35.0 | 38.0 | 47.5 | 42.35 | 34.6 | 37.0 | 47.0 | 40.00 | 34.1 | 35.8 | 45.0 | 40.69 | 34.6 | 36.0 | 44.6 |
| Inferior outer | 27.44 | 22.0 | 23.0 | 32.0 | 27.29 | 20.0 | 20.2 | 31.0 | 25.91 | 20.6 | 22.0 | 30.0 | 26.10 | 22.3 | 23.0 | 29.6 |

**Table 10.**
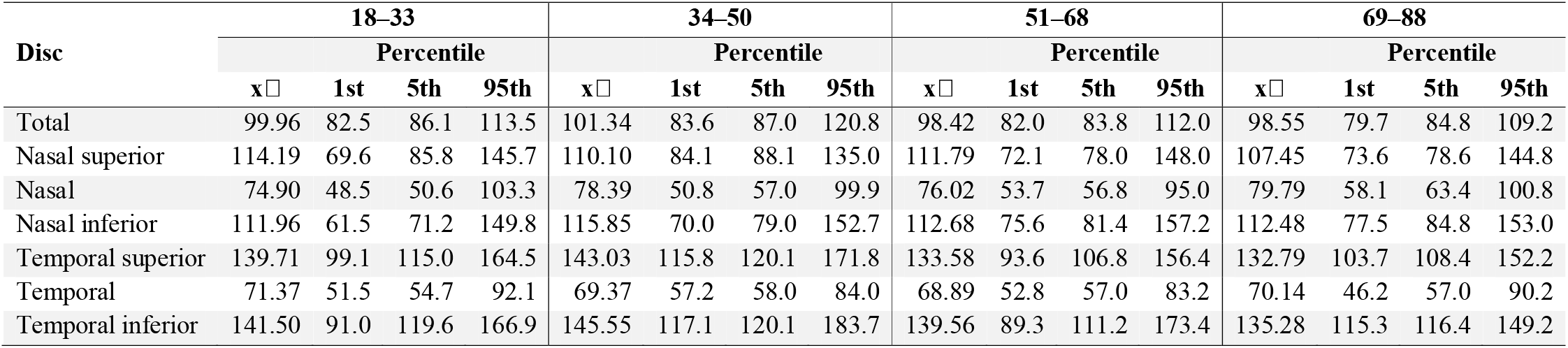
Retinal nerve fibre layer disc distribution by age group, showing mean, first, fifth and 95th percentile values for each segment.

Linear regression analysis showed a significant negative correlation between the total retinal volume and age (r = –0.2561), GCL volume and age (–0.2911) and IPL volume and age (– 0.3194). There was no significant correlation found between the RNFL and age. Figs 3 and 4 demonstrate trend lines for layer measurements against age. Table 11 demonstrates the regression analysis of segmental retinal thickness (RT) values against age. The strongest three significant negative associations were found in the superior inner IPL (r = –0.3444), nasal outer IPL (r = –0.3217) and inferior inner IPL (r = –0.3179) segments. The only significant positively correlated segment was the temporal inner RNFL (r = 0.1929). The only significant association between age and thickness at the peripapillary disc scan was the superior temporal sector (r = –0.1910) (Table 12).

**Table 11.** Regression analysis of layer thickness (µm) against age (years) and p-value for each macular segment.

| Segment | Retina |  | RNFL |  | GCL |  | IPL |  |
| --- | --- | --- | --- | --- | --- | --- | --- | --- |
|  | R | p | R | p | R | p | R | p |
| Volume | -0.2561 | 0.0003 | 0.0658 | 0.3544 | -0.2911 | <0.0001 | -0.3194 | <0.0001 |
| Centre | 0.0089 | 0.9008 | 0.0011 | 0.9875 | -0.1500 | 0.0340 | -0.0585 | 0.4103 |
| Nasal inner | -0.1821 | 0.0098 | -0.0003 | 0.9969 | -0.2821 | 0.0001 | -0.2930 | <0.0001 |
| Nasal outer | -0.3151 | <0.0001 | -0.0574 | 0.4191 | -0.3051 | <0.0001 | -0.3217 | <0.0001 |
| Superior inner | -0.1438 | 0.0421 | 0.0572 | 0.4214 | -0.2954 | <0.0001 | -0.3444 | <0.0001 |
| Superior outer | -0.2466 | 0.0004 | 0.0964 | 0.1743 | -0.2494 | 0.0004 | -0.2873 | <0.0001 |
| Temporal inner | -0.1117 | 0.1152 | 0.1929 | 0.0062 | -0.1847 | 0.0088 | -0.2071 | 0.0033 |
| Temporal outer | -0.1943 | 0.0058 | <0.0001 | 0.3556 | -0.2516 | 0.0003 | -0.2001 | 0.0045 |
| Inferior inner | -0.0532 | 0.4540 | <0.0001 | 0.9996 | -0.2841 | <0.0001 | -0.3179 | <0.0001 |
| Inferior outer | -0.2521 | 0.0003 | 0.0033 | 0.9627 | -0.2233 | 0.0015 | -0.2138 | 0.0024 |

**Table 12.**
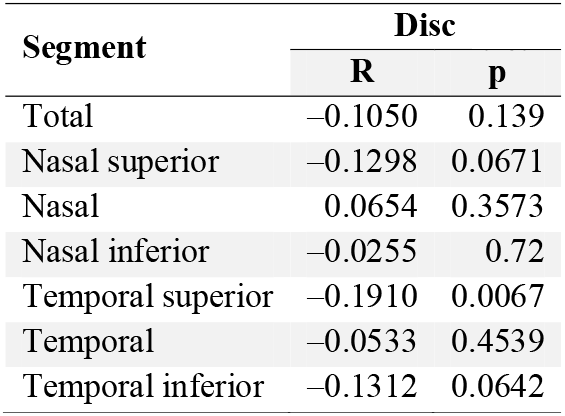
Regression analysis of layer thickness (µm) against age (years) and p-value for each disc segment.

**Fig 3.**
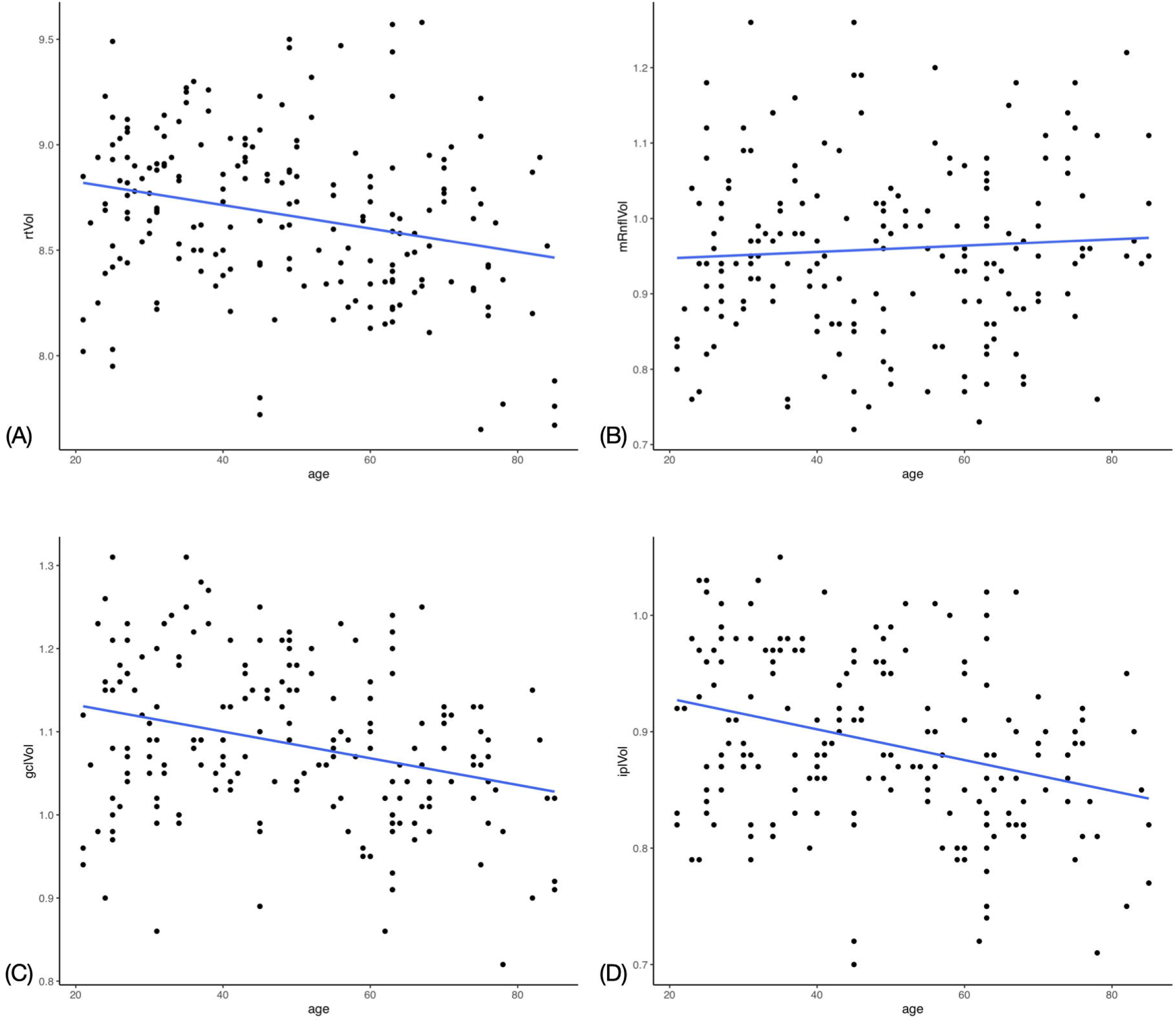
Scatter plot with trend line for macular optical coherence tomography measurements against age. (A) Total retinal volume. (B) Retinal nerve fibre layer volume. (C) Ganglion cell layer volume. (D) Inner plexiform layer volume.

**Fig 4.**
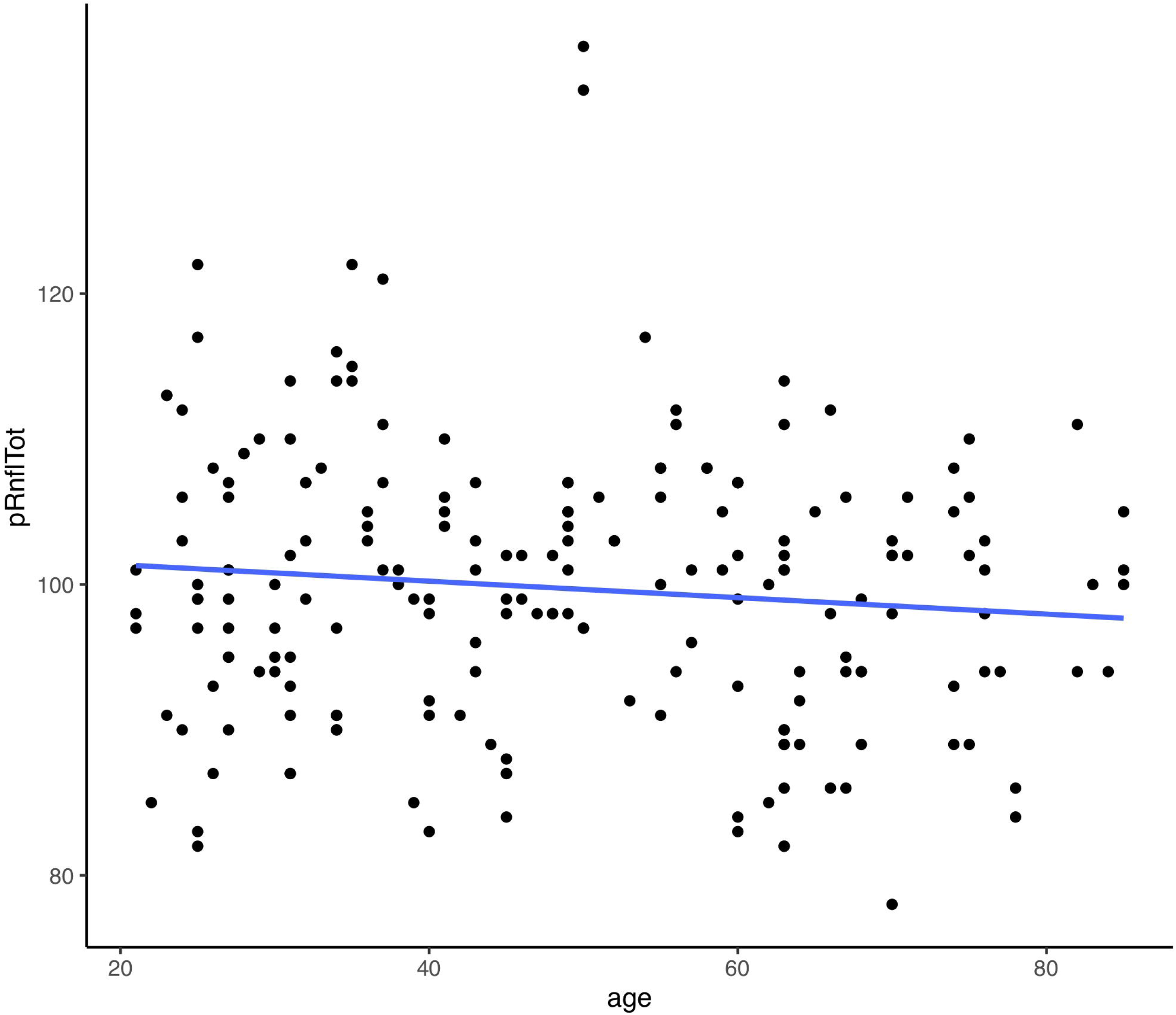
Scatter plot with trend line for mean peripapillary retinal nerve fibre layer thickness against age.

### Association between refractive error and thickness

Retinal thickness segments, except for the nasal outer, were not significantly correlated with refractive error. In order of magnitude, nasal outer (r = −0.2020), inferior outer (r = −0.1570) and inferior inner (r = −0.1314) RNFL segments were negatively correlated. Central RNFL was positively correlated (r = 0.1426). GCL segments were only negatively correlated; superior inner (r = −0.2034), nasal inner (r = −0.1782), temporal inner (r = −0.1534) and nasal outer (r = −0.1478). The IPL had the most segments significantly correlated with refractive error all of which were negative and included IPL volume (r = − 0.1668), nasal inner (r = −0.2154), nasal outer (r = −0.1946), superior inner (r = −0.2205), temporal inner (r = −0.1755), and inferior inner (r = −0.2448) (Table 13). Nasal (r = 0.1863) and nasal inferior (r = 0.1924) disc segments were significantly positively correlated while the temporal sector was negatively correlated (r = −0.2365).

**Table 13.**
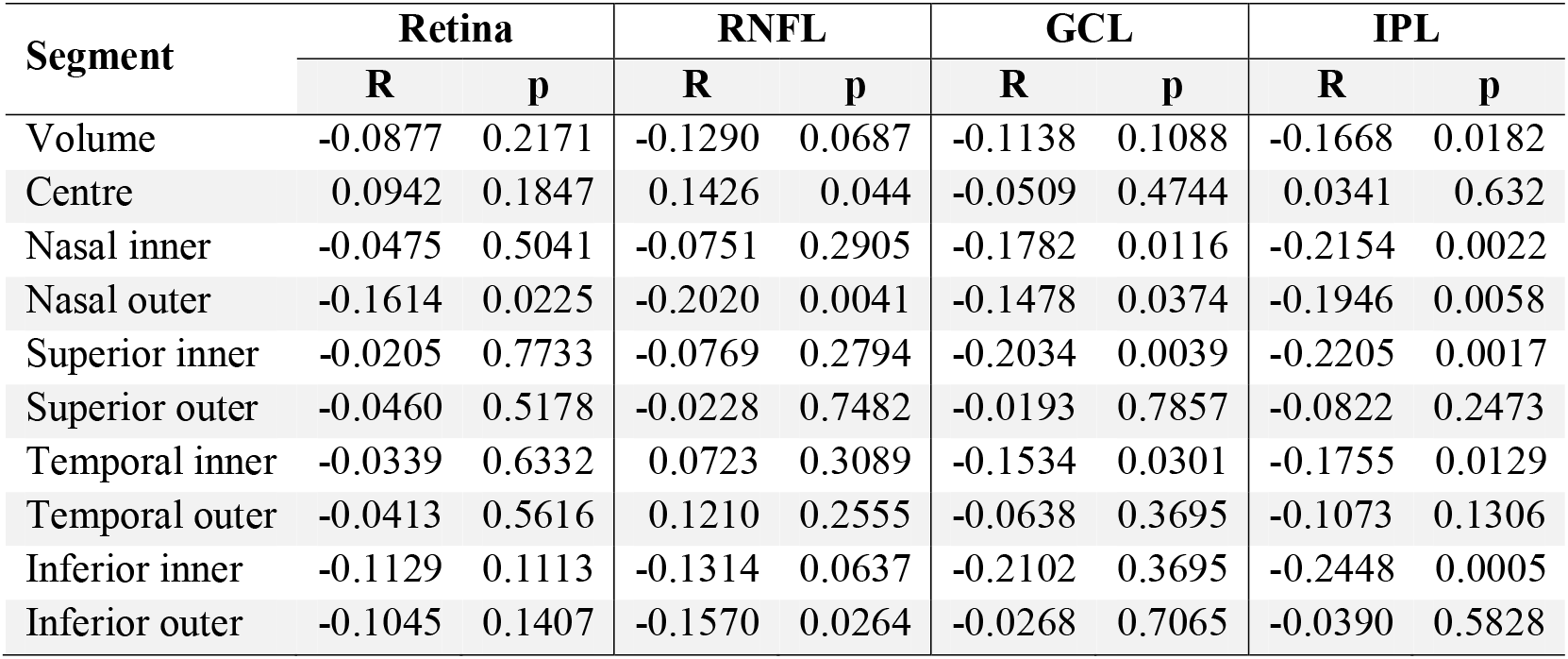
Regression analysis of layer thickness (µm) against refractive error (dioptres) and p-value for each macular segment.

**Table 14.** Regression analysis of layer thickness (µm) against refractive error (dioptres) and p-value for each disc segment.

| Segment | Disc |  |
| --- | --- | --- |
|  | R | p |
| Total | 0.0899 | 0.2056 |
| Nasal superior | 0.0383 | 0.5903 |
| Nasal | 0.1863 | 0.0083 |
| Nasal inferior | 0.1924 | 0.0063 |
| Temporal superior | -0.0722 | 0.3096 |
| Temporal | -0.2365 | 0.0007 |
| Temporal inferior | 0.1029 | 0.1472 |

## Discussion and Conclusion

SD OCT has revolutionised the diagnostic and prognostic capabilities for a number of ophthalmic conditions, including diabetic macular oedema, glaucoma, age-related macular degeneration and compressive optic neuropathy.[9,10] Normative databases provide the reference values against which to compare cases of disease states. Although several studies report the normative data for SD OCT,[11–13] to the author’s knowledge, no data exist for SD OCT that includes both macular GCL and peripapillary RNFL scans at the A-scan level. The A-scan is a single point of interrogation on the retina that represents the most basic unit of measurement data in the OCT. Equivalent A-scans represent the same anatomical location across patients. The comparison of A-scan measurements between patients is a more precise analysis than the standard ETDRS grid, which is based on averaged A-scan values across the segments. In the advent of analytical methodologies, such as artificial intelligence and machine learning, it is becoming increasingly possible to use larger volumes of data. The primary purpose of reporting normative A-scans is to facilitate further research into patterns of disease development at the retina beyond the confines of standardised grids, such as ETDRS, using such methodologies.

In the creation of a normative database, inclusion and exclusion criteria must be carefully considered to realistically represent the distribution of normal and diseased populations of interest. This study aimed to create a normative dataset of OCT scans for ongoing research of glaucoma and compressive optic neuropathies within the metropolitan region of Melbourne, Australia. An ethnically diverse population was used to represent the target population. All nine of the broad ethnic groups present in Australia were represented in our study population in similar proportions.

Five groups showed no statistical difference compared to the known proportions in Melbourne. Given the small sample size and large number of ethnic groups, the database was representative of the target population. The ethnic groups used in this study were extracted from population census data that were designed according to political and geographical boundaries. This may have caused the grouping of some participants with differing genetic influences on their retinal structure. For example, the Americas ethnic group included both African American and Caucasian American people that have been shown to have different OCT measurements.[14,15] This was an unavoidable limitation set by the Australian census groups.[16]

This study found a negative association between age and macular thickness, which has been previously demonstrated. All the sectors of the ETDRS grid have been reported to be negatively associated with increasing age, except for the central 1 mm of the retina.[12,17–19] This study is consistent with these findings; however, the negative correlation found in the temporal inner and inferior inner segments was found not to be statistically significant. Similar to other studies, the central area had almost no association (r = 0.00899, p = 0.9008). This study identified that the GCL and IPL thickness and volume for all the sectors (excluding the centre for the IPL) decreased with age. This was in keeping with findings from previous studies.[12,17] This finding represents a normal process of ageing with loss of RNFLs over time, which has been reported to be between 0.3% and 0.6% of neurons lost per year.[20] This is important for differentiating losses due to age-related processes from those caused by disease processes, such as glaucoma or compressive optic neuropathy.

A statistically significant positive correlation was found between increasing age and temporal inner RNFL thickness (r = 0.1929, p = 0.0062). Similarly, Nieves-Moreno et al. found a positive correlation between temporal inner RNFL and age (r = 0.256, p = <0.001).^11^ No other significant correlations were found between RNFL volume or thickness and age in this study. Given that the temporal inner is the thinnest RNFL sector, this study postulated that it would be proportionally the most affected by an increasing thickness observed in the internal limiting membrane with age and, therefore, correlate positively with age. Males were found to have a significantly higher RT, RNFL, GCL and IPL thickness than females. This was most evident in the inner layers, which is similar to the OCT results reported in other studies.[12,17]

The Limitations of this study include its retrospective design, which led to missing variables that may have affected the OCT measurements, including the axial length and refraction. This study provides normal baseline data for the comparison of various macular diseases. These data will also be used to monitor patients with glaucoma and compressive optic neuropathy at The RMH.

## Data Availability

Data will be made available after publication

## Acknowledgements

Amanda Rebbechi – medial photographer; Joss Dimock – medical photographer, Debbie Curran – medical photographer;

